# Omicron (BA.1) SARS-CoV-2 variant is associated with reduced risk of hospitalization and length of stay compared with Delta (B.1.617.2)

**DOI:** 10.1101/2022.01.20.22269406

**Authors:** André Peralta-Santos, Eduardo Freire Rodrigues, Joana Moreno, Vasco Ricoca, Pedro Casaca, Eugenia Fernandes, João Paulo Gomes, Rita Ferreira, Joana Isidro, Miguel Pinto, Vítor Borges, Luís Vieira, Sílvia Duarte, Carlos Sousa, José Pedro Almeida, Luís Menezes, Bibiana I. Ferreira, Ana Matias, Ana Pelerito, Samanta Freire, Teresa Grilo, Cláudia Medeiros Borges, Vera Moutinho, Andreia Leite, Irina Kislaya, Ana Paula Rodrigues, Pedro Pinto Leite, Baltazar Nunes

## Abstract

**Introduction:** Early reports showed that Omicron (BA.1) SARS-CoV-2 could be less severe. However, the magnitude of risk reduction of hospitalization and mortality of Omicron (BA.1) infections compared with Delta (B.1.617.2) is not yet clear. This study compares the risk of severe disease among patients infected with the Omicron (BA.1) variant with patients infected with Delta (B.1.617.2) variant in Portugal.

**Methods:** We conducted a cohort study in individuals diagnosed with SARS-CoV-2 infection between 1^st^ and 29^th^ December 2021. Cases were individuals with a positive PCR test notified to the national surveillance system. SARS-CoV-2 variants were classified first by whole genomic sequencing (WGS) and, if this information was unavailable, by detecting the S gene target failure. We considered a hospitalization for all the patients admitted within the 14 days after the SARS-CoV-2 infection; after that period, they were censored.

The comparison of the risk of hospitalization between Omicron (BA.1) and Delta (B.1.617.2) VOC was estimated using a Cox proportional hazards model. The mean length of stay was compared using linear regression, and the risk of death between Omicron and Delta patients was estimated with a penalized logistic regression. All models were adjusted for sex, age, previous infection, and vaccination status.

**Results:** We included 15 978 participants aged 16 or more years old, 9 397 infected by Delta (B.1.617.2) and 6 581 infected with Omicron (BA.1). Within the Delta (B.1.617.2) group, 148 (1.6%) were hospitalized, and 16 (0.2%) were with the Omicron (BA.1). A total of 26 deaths were reported, all in participants with Delta (B.1.617.2) infection. Adjusted HR for hospitalization for the Omicron (BA.1) variant compared with Delta (B.1.617.2) was 0.25 (95%CI 0.15 to 0.43). The length of stay in hospital for Omicron (BA.1) patients was significantly shorter than for Delta (confounding-adjusted difference -4.0 days (95%CI -7.2 to -0.8). The odds of death were 0.14 (95% CI 0.0011 to 1.12), representing a reduction in the risk of death of 86% when infected with Omicron (BA.1) compared with Delta (B.1.617.2).

**Conclusion:** Omicron (BA.1) was associated with a 75% risk reduction of hospitalization compared with Delta (B.1.617.2) and reduced length of hospital stay.

## Background

The Omicron (BA.1) SARS-CoV-2 has been a variant of concern (VOC) since late November 2020 (1) and swept South Africa, Europe and other regions, becoming rapidly dominant. The Omicron (BA.1) has mutations associated with increased transmissibility, immune escape, and a higher risk of previous infection (2, HYPERLINK \l "bookmark2" 3). It also harbors a deletion (spike Δ69–70) leading to “S gene target failure” (SGTF) by some real-time PCR assays (e.g. TaqPath COVID-19, ThermoFisher, Waltham, MA, United States). This distinctive feature compared with the Delta (B.1.617.2) can be used as a marker (4), as it has been done previously with the Alpha variant (5).

The first real-world studies in England (6), Scotland (7) and Denmark (8) found reduced vaccine effectiveness for symptomatic Omicron (BA.1) infections for both complete vaccination schemes and booster doses. Although studies about vaccine effectiveness for symptomatic are essential, severity studies are complementary and can inform about the potential impact on hospitalizations and COVID-19 death for Omicron.

Early reports showed that Omicron could be less severe (9). In animal models, the Omicron showed less severe symptoms (10) and less viral load in the lungs; in vitro T cell response was also preserved with the Omicron variant (11). In real-world human studies, reduced severity was confirmed in studies in Scotland (7), England (6, 12), Canada(13) and the US (California)(14); while studies consistently showed a risk reduction the degree of risk reduction (from 80% to 45%) of hospitalization varied considerably. Furthermore, length of stay and risk of death resulting from Omicron cases have not been described.

We aim to study the risk of hospitalization and death from Omicron (BA.1) infections compared with Delta (B.1.617.2) variant infections in a population with high vaccination coverage, and assess differences in viral loads and length of stay in hospitalizations, following similar studies done for Alpha and Delta variant (15).

## Methods

### Study design and population

We designed a cohort study to assess the risk of hospitalization and death of the individuals infected with Omicron (BA.1) and Delta (B.1.617.2) variants. The study population was individuals eligible for vaccination (>16 years old) diagnosed with SARS-CoV-2 infection/COVID-19 by nasopharyngeal swab tested with RT-PCR from December 1^st^ and 29^th^ notified through the laboratory service of the existing surveillance system (SINAVE), in Portugal mainland. We included individuals with samples classified either by whole-genome sequencing or Spike Gene Target Failure (SGTF). Both symptomatic and asymptomatic individuals were included.

### Sample collection and variant classification

We used two different methods to identify which cases were Delta (B.1.617.2) or Omicron (BA.1) variants infections. We used information from a nationwide network group of laboratories (UNILABS, ABC and CVP) that performs RT-PCR tests for SARS-CoV-2 using Thermofisher TaqPath assay, targeting three regions of the SARS-CoV-2 genome: ORF1ab, N and S genes. Their samples are classified as BA.1 (no amplification) or B.1.617.2 (amplification), according to SGTF status. Only samples having both N and ORF1a positive signals and Ct values <=30 were considered. Secondly, we use Whole-Genome Sequencing (WGS) data provided by the National Health Institute (INSA) that routinely performs sequencing on random samples notified to SINAVE. Variants were classified first by whole genomic sequencing (WGS) and, if this information was unavailable, by detecting the S gene target failure (SGTF) (16).

### Outcomes: hospitalization and death

We defined COVID-19 hospitalization as any admission to a public hospital in Portugal mainland within the 14 days following a positive sample SARS-CoV-2 collection, notified in the surveillance system. Admission data was obtained through the Central Hospital Morbidity Database and Integrated Hospital Information System. The Central Hospital Morbidity Database gathers data from all public hospitals in Portugal mainland, which account for most hospitals in Portugal and most hospitals admitting patients with COVID-19. Individuals with a SARS-CoV-2 diagnosis (by sample collection day) or notification after the admission date were excluded.

A COVID-19 death was defined as any record of death on the national Death Certificate Information System (SICO) with COVID-19 as the primary cause of death (ICD-10 *code U*.*071*) according to the WHO classification (17). SICO platform allows the issuance of a death certificate for each person who dies in Portugal (18). A COVID-19 death was defined as any record of death on the national Death Certificate Information System (SICO) with COVID-19 as the primary cause of death (ICD-10 *code U*.*071*) according to the WHO classification (17). SICO platform allows the issuance of a death certificate for each person who dies in Portugal (18).

### Vaccination status and demographic covariates

We obtained COVID-19 vaccination status through the electronic national vaccination register (VACINAS). Vaccination exposure was indexed at the date of COVID-19 diagnosis and classified as: (i) unvaccinated: no register of vaccine administration prior to diagnosis; (ii) incomplete vaccination: SARS-CoV-2 infection/COVID-19 diagnosis anytime between the uptake of the first dose of a COVID-19 vaccine (ChAdOx1, BNT162b2 or mRNA-1273) and up to 14 days after the second dose uptake; (iii) complete vaccination: 14 or more days following second dose vaccine uptake or first dose of Ad26.COV2.S or up to 14 days after the booster dose uptake; and (iv) complete vaccination plus booster: 14 or more days following booster dose vaccine uptake. Information about age, sex and date of diagnosis was routinely collected on surveillance system SINAVE and was extracted from it. Previous infection was defined as a PCR or rapid antigen SARS-CoV-2 notification for the same individual more than 90 days apart.

### Data source, record-linkage, and follow-up

We used data from different sources: databases from the three laboratories with information about unique identifiers (national health service number), sample collection date, Ct value and SGTF classification; this was combined (full join) with data from sequencing was provided by the National Institute of Health Dr Ricardo Jorge. Duplicates were excluded using the unique identifier. Afterward, we linked (left join) the former database with SINAVE, the national epidemiological and laboratory surveillance platform for SARS-CoV-2 infections, providing age and sex information. SICO, the electronic death certificate platform, provided information about COVID-19 basic cause of death and date of death. VACINAS, the electronic vaccination platform, provided information about vaccination status. SONHO, the hospital admission platform of the National Health System, provided data about hospitalization. Data from the laboratories were extracted on December 29 and followed up (hospitalization and death) until January 11, 2022.

### Statistical Analysis

We compared characteristics of SARS-CoV-2 cases using central tendency and dispersion measures for continuous variables and absolute and relative frequencies for categorical variables. To compare the mean, we used the T-test and the Chi-squared test for relative frequencies.

The confounding-adjusted hazard ratio of hospitalization, with 95% confidence intervals (CI), was calculated using a proportional hazards Cox model, adjusted for sex, age, previous infection and vaccination status (equation 1).

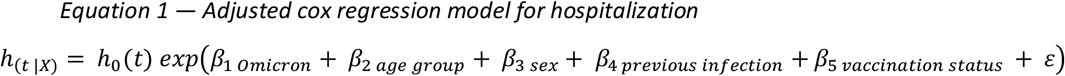

Where *h*_(*t*|*X*)_ is the hazard at the time t of the outcome hospitalization conditional on covariates X (Omicron, age group, sex, previous infection and vaccine status), *h*_0_(*t*) is the baseline hazard for the outcome, exp (*β*_1 *Omicron*_) the hazard ratio for the outcome being infected with Omicron variant compared with Delta, exp (*β*_2 *age group*_) are the hazard ratios for the outcome being 80 years or older, 65 years old to 80 compared with being 16 to 65 years old, exp (*β*_3 *sex*_) the hazard ratio for the outcome being a female compared with being male, exp (*β*_4 *previous infection*_) is the hazard ratio for the outcome of primary infection compared with having a previous infection, exp (*β*_5 *vaccination status*_) is the hazard ratio for the outcome having incomplete, complete, or complete plus booster compared with unvaccinated, *ε* is the error term.

We tested the proportionality of risk assumptions using visual inspection of scaled Schoenfeld residuals (19). We used the R survival package to fit the proportional hazards Cox model (20). Due to the nature of our analysis assumed a type I right censoring, thus using a fixed end of follow-up date defined as 14 days after the sample collection date for non-hospitalized or the day of hospitalization. Analyses were based on a complete case analysis.

We calculated the length of stay as the difference between discharge and admission dates. We used a linear regression model to calculate the confounding-adjusted difference in length of stay between Omicron (BA.1) and Delta (B.1.617.2), adjusted for sex, age, previous infection, and vaccination status. We also performed a stratified analysis by vaccination status in the adjusted model.

The risk of death was assessed using logistic regression adjusted for sex, age, previous infection, and vaccination status. To account for the fact that we did not observe deaths in Omicron study participants, we used a penalized maximum likelihood logistic regression (21) (supplementary material, equation S1).

All tests were two-sided, and a P-value < .05 was considered statistically significant. All statistical analyses were performed on R® using R Studio version 1.4 (22).

### Sensitivity Analysis

As an additional analysis to account for potential biases regarding the week of diagnosis, we performed a matching based on the week and age group of SARS-CoV-2 infection diagnosis using variable ratio matching, at a ratio of 1 to k, where k is the maximum number of B.1.617.2 cases available on the respective week. We used a nearest-neighbor algorithm with the “Matchit”(23) CRAN package.

### Ethical Statement

The genomic surveillance of SARS-CoV-2 in Portugal is regulated by the Assistant Secretary of State and Health Executive Order (Despacho n.° 331/2021 of January 11, 2021). The research on genomic epidemiology of SARS-COV-2 received the clearance of the Ethics Committee of INSA on March 30, 2021.

## Results

### Study participants characteristics

From 1^st^ to 29^th^ December 2021, we identified through WGS or SGTF with successful record linkage 22 960 SARS-CoV-2 cases in Portugal considering the inclusion criteria. 6 982 (30%) were excluded due to missing information about previous infection. Hence the study included 15 978 participants, 9 397 infected with Delta (B.1.617.2) and 6 581 with Omicron (BA.1) variant.

Study participants with Omicron (BA.1) variant were on average younger, 37.1(SD=14.8) for Omicron cases and 43.4 (SD=15.8) years for Delta (B.1.617.2). Participants with Omicron (BA.1) were more frequently female (51.1% versus 49.3%) and with complete plus booster vaccination status (4.5% versus 2.1%). Also, previous infection in participants with Omicron (BA.1) was four times higher than Delta (B.1.617.2), 6.8% versus 1.6%. Patient characteristics at baseline were not balanced between the groups for some potential confounders (Table 1 and Supplementary Materials figure S1).

**Table 1.**
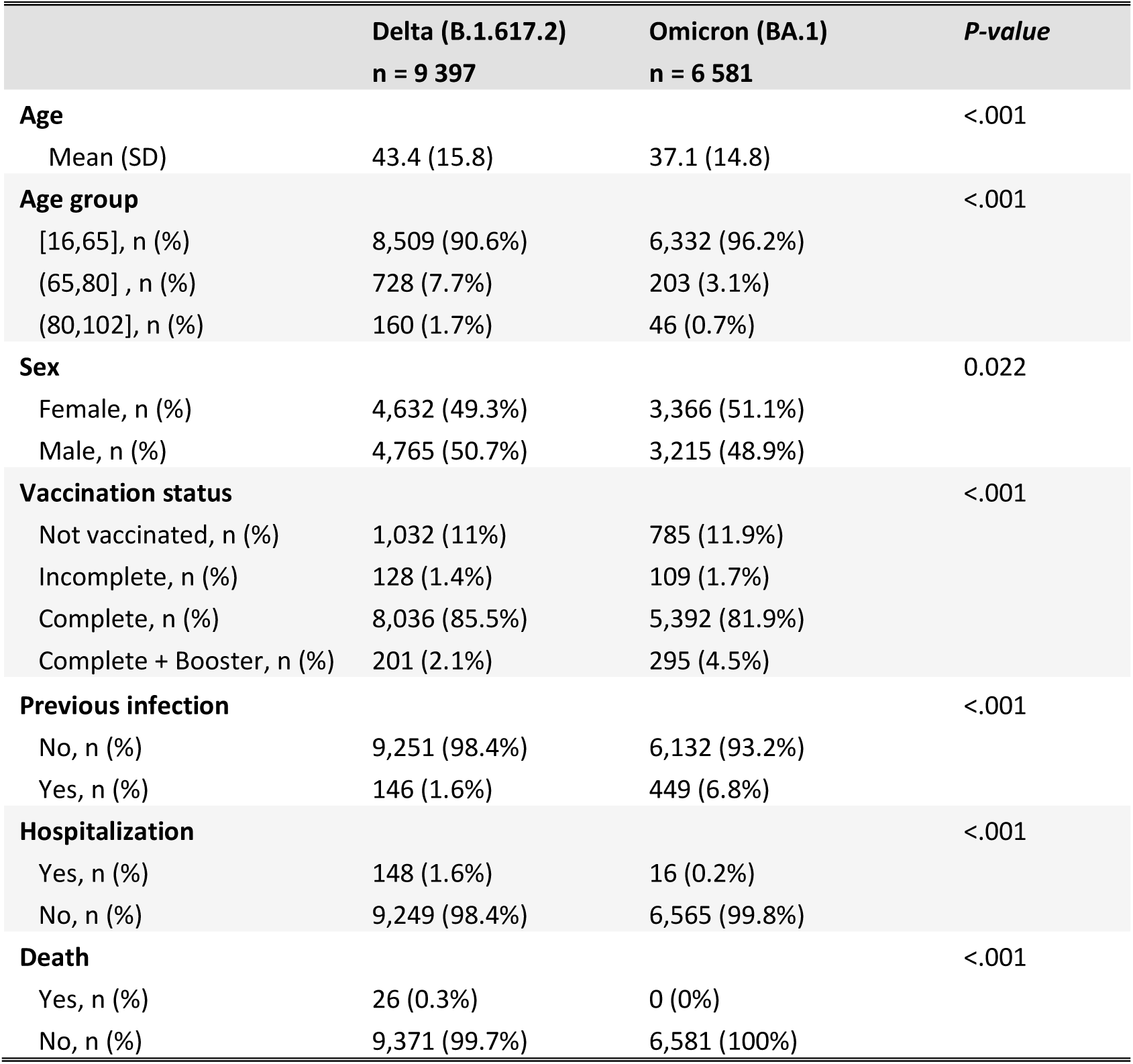
Sociodemographic and clinical characteristics of the study sample, according to the identified SARS-CoV-2 variant

|  | Delta (B.1.617.2)<br>n = 9 397 | Omicron (BA.1)<br>n = 6 581 | P-value |
| --- | --- | --- | --- |
| **Age** |  |  | <.001 |
| Mean (SD) | 43.4 (15.8) | 37.1 (14.8) |  |
| **Age group** |  |  | <.001 |
| [16,65], n (%) | 8,509 (90.6%) | 6,332 (96.2%) |  |
| (65,80] , n (%) | 728 (7.7%) | 203 (3.1%) |  |
| (80,102], n (%) | 160 (1.7%) | 46 (0.7%) |  |
| **Sex** |  |  | 0.022 |
| Female, n (%) | 4,632 (49.3%) | 3,366 (51.1%) |  |
| Male, n (%) | 4,765 (50.7%) | 3,215 (48.9%) |  |
| **Vaccination status** |  |  | <.001 |
| Not vaccinated, n (%) | 1,032 (11%) | 785 (11.9%) |  |
| Incomplete, n (%) | 128 (1.4%) | 109 (1.7%) |  |
| Complete, n (%) | 8,036 (85.5%) | 5,392 (81.9%) |  |
| Complete + Booster, n (%) | 201 (2.1%) | 295 (4.5%) |  |
| **Previous infection** |  |  | <.001 |
| No, n (%) | 9,251 (98.4%) | 6,132 (93.2%) |  |
| Yes, n (%) | 146 (1.6%) | 449 (6.8%) |  |
| **Hospitalization** |  |  | <.001 |
| Yes, n (%) | 148 (1.6%) | 16 (0.2%) |  |
| No, n (%) | 9,249 (98.4%) | 6,565 (99.8%) |  |
| **Death** |  |  | <.001 |
| Yes, n (%) | 26 (0.3%) | 0 (0%) |  |
| No, n (%) | 9,371 (99.7%) | 6,581 (100%) |  |

### Outcomes

We observed 164 hospitalizations and 26 deaths between 1^st^ and 29^th^ December. There were 148 (1.6%) hospitalizations in participants with Delta variant and 16 (0.2%) with Omicron (BA.1). All the 26 deaths were reported in participants with Delta (B.1.617.2) variant.

For hospitalized participants, the mean time from sample collection to hospitalization was 3.4 days (SD=3.7) for Delta (B.1.617.2) participants and 5.0 days (SD=3.9) for Omicron (BA.1). For the outcome of death, the mean time from diagnosis to death was for Delta (B.1.617.2) participants and 9.1 days (SD=5.3).

We observed no Intensive Care Unit (ICU) admission for Omicron (BA.1) cases; 17 patients were (11.5%) admitted to ICU for Delta (B.1.617.2) cases. The mean length of stay was also higher for patients with Delta (B.1.617.2), 8.6 days (SD=5.9) and 3.7 days (SD=2.9) for Omicron (BA.1), see table 2.

**Table 2.** Characteristics of COVID-19 hospitalized patients

|  | Delta (B.1.617.2)<br>n = 148 | Omicron (BA.1)<br>n = 16 | P-value |
| --- | --- | --- | --- |
| **Time to hospitalization** |  |  | 0.001 |
| Mean days (SD) | 3.4 (3.7) | 5.0 (3.9) |  |
| **ICU admission** |  |  | 0.317 |
| No | 131 (88.5%) | 16 (100%) |  |
| Yes | 17 (11.5%) | 0 (0%) |  |
| **Length of stay (days)<sup>1</sup>** |  |  |  |
| Mean (SD) | 8.6 (5.9) | 3.7 (2.9) |  |
| Diff. |  | -4.9 (95%CI -7.9 to -1.8) | 0.002 |
| Conf. Adj. Diff. |  | -4.0 (95%CI -7.2 to -0.8) | 0.02 |
| **Ct values<sup>2</sup>** |  |  |  |
| N Mean (SD) | 18.6 (4.9) | 19.3 (4.6) | 0.639 |
| ORF1ab Mean (SD) | 18.1 (4.8) | 18.1 (3.6) | 0.993 |

ICU – Intensive care unit dedicated to COVID-19
1. Based on 132 patients for Delta and 15 patients for Omicron, 2. Based on 63 patients for Delta and 13 for Omicron Diff. Unadjusted difference | Confounding-Adj. Diff. The adjusted difference for age, sex, previous infection, and vaccination status using linear regression

**Table 3.** Death odds ratio between Omicron and Delta

| Omicron VS Delta | Odds Ratio | Lower CI | Upper CI | P-value |
| --- | --- | --- | --- | --- |
| **Death** |  |  |  |  |
| Unadjusted | 0.026 | 0.00021 | 0.191 | <0.001 |
| Adjusted* | 0.14 | 0.0011 | 1.12 | 0.06 |

\*The adjusted for age, sex, previous infection, and vaccination status

### Hospitalization

The crude hazard ratio of hospitalization was 0.15 (95%CI 0.091 to 0.25) for Omicron compared with Delta. The age, sex, previous infection and, vaccination adjusted HR for hospitalization for the Omicron (BA.1) variant compared with Delta (B.1.617.2) was 0.25 (95%CI 0.15 to 0.43), figure 1. Hence the Omicron (BA.1) variant cases had a 75% risk reduction of hospitalization compared with Delta (B.1.617.2) (95%CI 57% to 85%). We report no statistically significant difference in the Ct values between Omicron (BA.1) and Delta (B.1.617.2) in hospitalized participants. The confounding-adjusted difference in length of stay of -4.0 days (95%CI -7.2 to -0.8), between Delta and Omicron is not explained by higher viral loads, measured as Ct Values at the time of diagnosis. In the stratified analysis, in the unvaccinated strata, the adjusted HR for hospitalization for the Omicron (BA.1) variant compared with B.1.617.2 was 0.21 (95%CI 0.08 to 0.51). In the vaccinated strata (complete and complete+booster), the adjusted HR for hospitalization for the Omicron (BA.1) variant compared with Delta (B.1.617.2) was 0.34 (95%CI 0.13 to 0.87), see supplement materials, figures S2 and S2. We found no significant violation of the proportionality of hazards assumptions (see supplementary material, figure S4).

**Figure 1.**
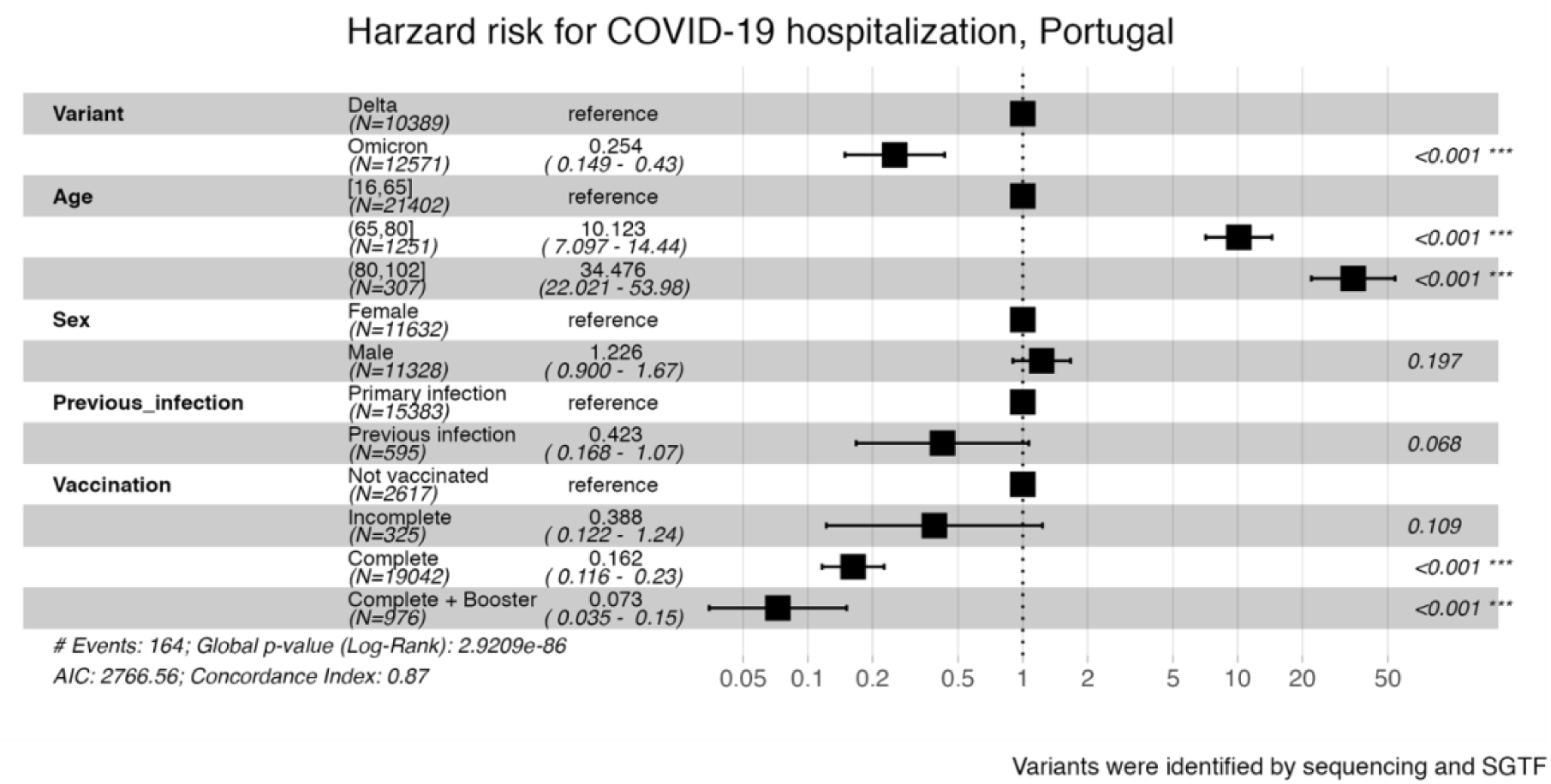
The hazard ratio for hospitalization in the fully adjusted cox model

### COVID-19 death

All the 26 study participants who died were Delta (B.1.617.2) cases. The average age at the time of death was 83.1 (SD=8.9). In the group of participants that died, 20 (77%) were hospitalized, and no one had previous infection. Regarding vaccination status, 8 (31%) were not vaccinated, 16 (62%) had complete vaccination, and two complete boosters (8%). In the unadjusted model, the odds ratio of death was 0.026 (95% CI 0.00021 to 0.191), comparing patients infected with Omicron (BA.1) cases with patients infected with Delta (B.1.617.2). However, after adjusting for age, sex, vaccination, and previous infection, the odds ratio of death was 0.14 (95% CI 0.0011 to 1.12), see supplement materials, table S1. That represents a reduction in the risk of death of 86% when infected with Omicron (BA.1) compared with Delta (B.1.617.2), although with a wide confidence interval.

### Sensitivity Analysis

After we matched for the week and age group of diagnosis, the study sample was 10 941 individuals, and in the adjusted HR for hospitalization for the Omicron (BA.1) variant compared with B.1.617.2 was 0.24 (95%CI 0.11 to 0.50) times the risk of Delta (B.1.617.2).

## Discussion

We report a 75% risk reduction of hospitalization for participants infected with Omicron (BA.1) compared with Delta (B.1.617.2). The length of stay for hospitalized study Omicron (BA.1) participants was significantly shorter than for Delta (B.1.617.2) participants, mean difference -4.0 days (95%CI -7.2 to -0.8). However, there were no differences in mean Ct values between Omicron and Delta hospitalized participants. The risk of death was also lower; we show a reduction in the odds of death of 86% when infected with Omicron (BA.1) compared with Delta (B.1.617.2), OR 0.14 (95% CI 0.0011 to 1.12); however, the effect was highly uncertain.

Early assessment of the risk of hospitalization in South Africa founded an odds ratio (aOR) 0.2, 95% confidence interval (CI) 0.1-0.3) (9), adjusted for age, sex, previous infection, vaccination and comorbidities. In the Scottish study, the adjusted observed/expected ratio was 0.32 (95% CI 0.19, 0.52) for Omicron (BA.1) compared with Delta (B.1.617.2) (7). The Canadian study reports that the risk of hospitalization or death was 65% lower (hazard ratio, HR=0.35, 95%CI: 0.26, 0.46) among Omicron (BA.1) cases compared to Delta (B.1.617.2) cases (13). The study in California reports an adjusted hazard ratio for hospital admission associated with Omicron (BA.1) variant infection compared with Delta (B.1.617.2) of 0.48 (0.36-0.64) (14). Finally, the English study (12) reports an adjusted HR for hospitalization of 0.55 (0.51-0.59) for Omicron (BA.1) compared with Delta (B.1.617.2) (*“All Pillar 1 and Pillar 2 cases, ECDS Admitted”*). All these studies have different designs, but they were consistent with a risk reduction for hospitalized individuals infected with Omicron compared with Delta. Our study has a confounding-adjusted HR lower than most studies: HR 0.25 (95%CI 0.15 to 0.43); this could be due to residual confounding, the study participants with Omicron (BA.1) were younger than those with Delta (B.1.617.2), and we do not have data on comorbidities to adjust for. However, even after matching for the week of diagnosis and age group, the results are consistent with a risk reduction of a similar magnitude. In our study, the reduction in the risk of hospitalization for Omicron (BA.1) participants is maintained for not vaccinated individuals and vaccinated individuals, accounting for confounders. This result is consistent with the study from England (12).

It is also important to highlight that those infected with Omicron (BA.1) had a shorter length of hospitalization than Delta(B.1.617.2), mean difference of -4.0 days (95%CI -7.2 to -0.8). The study in California also reported a median duration of hospital stay of 3.4 (2.8-4.1) days shorter for hospitalized cases with Omicron variant infections than hospitalized patients with Delta variant infections (14). This fact has important implications in the management of hospital services and will reduce the impact of omicron waves on health systems, and to inform statistical models.

Our results also indicate a reduction in the risk of death of participants infected with Omicron, although with high uncertainty. Our study might be underpowered to study this risk reduction, but a reduction in the risk of death is plausible given the animal models showing less severe inflammation in the lungs (10), the preserved T cell function in vitro (11) and a reduced risk in hospitalizations in humans.

Our study has several strengths,1) we used an established method to identify Delta and Omicron variants, the SGTF and genome sequencing, 2) we also provided data from record linkage of study participants that were admitted to the hospital within 14 days of the diagnosis of the infection, that was used before (7), 3) we provide data from a short period (4 weeks), minimizing the probability of changes in the testing policies, clinical guidelines; 4) we adjusted for the main confounders associated with the probability of severe disease, especially vaccination and previous infection that could differ across time and variant.

Our study also had limitations; we did not have access to previous comorbidities, which could be an unaccounted confounder. Also, the study participants with Omicron and Delta had baseline differences that could influence the risk of hospitalization; although this was accounted for in the adjusted models, residual confounding could still be present. The adjusted risk increased the HR by 66%, hence the protection effect of omicron could be partially explained by age, sex, previous infection, and vaccination status.

Finally, we did not account for correcting hazard ratio estimates in unvaccinated individuals for under ascertainment of past infection status; that method was shown in the English study to decrease the risk reduction. Hence, we could be overestimating the risk reduction.

## Conclusion

Omicron was associated with a 75% risk reduction of hospitalization compared with Delta (B.1.617.2); the risk reduction is maintained for unvaccinated and vaccinated participants. Additionally, Omicron was associated with a shorter length of hospital stay, mean difference -4.0 days (95%CI -7.2 to -0.8), and reduced risk of death, however the latter with high uncertainty.

## Data Availability

All data produced in the present study are available upon reasonable request to the authors

