## Supplementary material for "Omicron (BA.1) SARS-CoV-2 variant is associated with reduced risk of hospitalization and length of stay compared with Delta (B.1.617.2)"

**Supplementary Materials**

*Equation S1 — Adjusted penalized logistic regression.*

$$Y_{(Death\ |X)} = \beta_0 + \beta_{1\ Omicron} + \beta_{2\ age\ group} + \beta_{3\ sex} + \beta_{4\ previous\ infection} + \beta_{5\ vaccination\ status} + \varepsilon$$

Where $Y_{(Death\ |X)}$ is the Odds of the outcome hospitalization conditional on covariates X (Omicron, age group, sex, previous infection and vaccine status), $\exp(\beta_{1\ Omicron})$ the odds ratio for the outcome being infected with Omicron variant compared with Delta, $\exp(\beta_{2\ age\ group})$ are the odds ratios for the outcome being 80 years or older, 65 years old to 80 compared with being 16 to 65 years old, $\exp(\beta_{3\ sex})$ the odds ratio for the outcome being a female compared with being male, $\exp(\beta_{4\ previous\ infection})$ is the odds ratio for the outcome of primary infection compared with previous infection, $\exp(\beta_{5\ vaccination\ status})$ is the odds ratio for the outcome having incomplete, complete, or complete plus booster compared with unvaccinated, $\varepsilon$ is the error term.

*Figure S1 — Age density per virus variant of the study population*

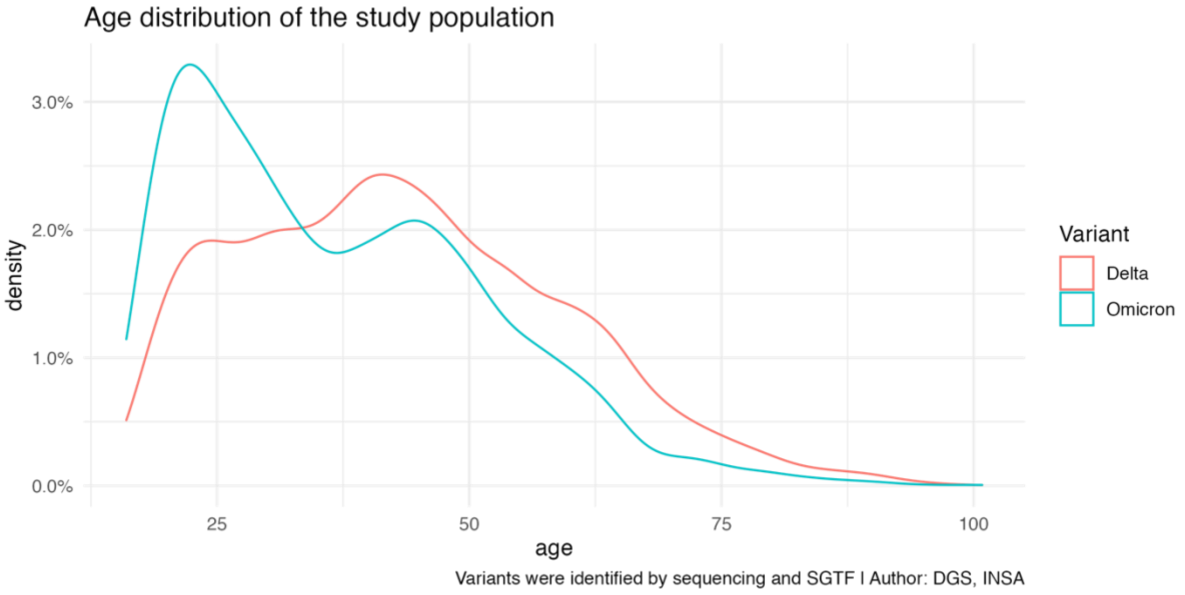

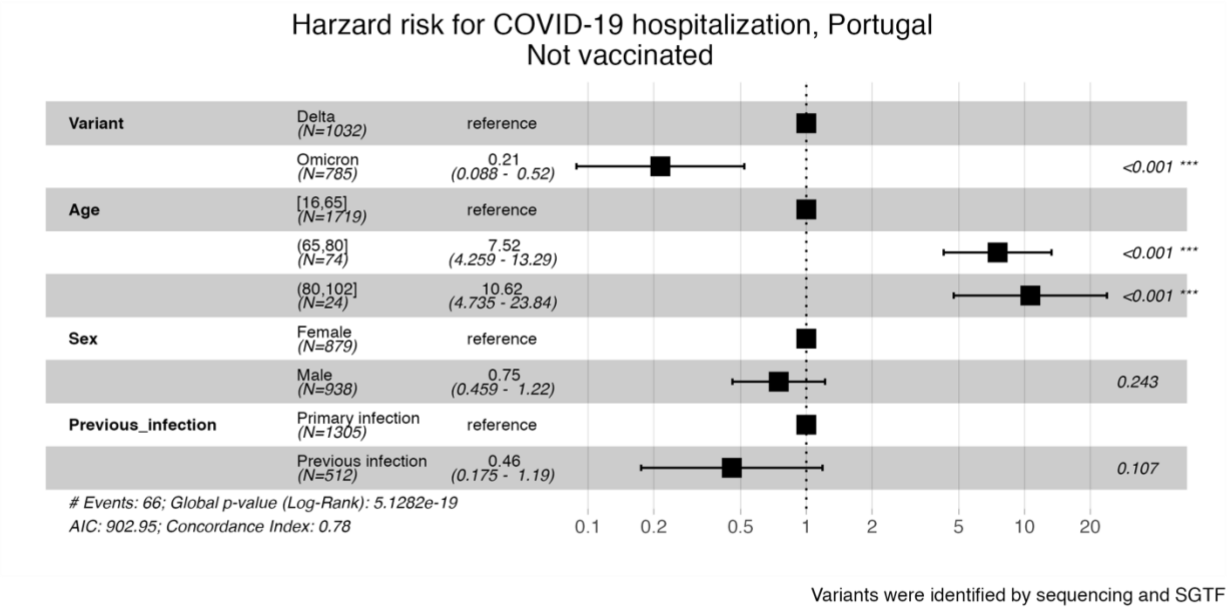


*Figure S2. Hazard Ratio for hospitalization in the adjusted model stratified for not vaccinated individuals*

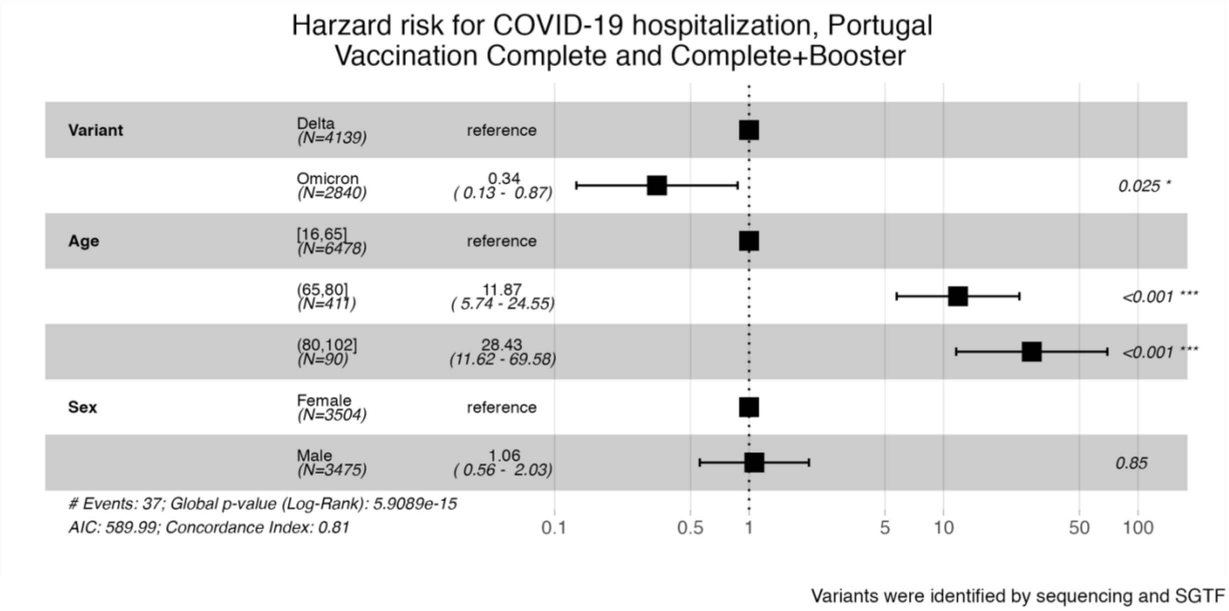


*Figure S3. Hazard Ratio for hospitalization in the adjusted model stratified for vaccinated individuals*

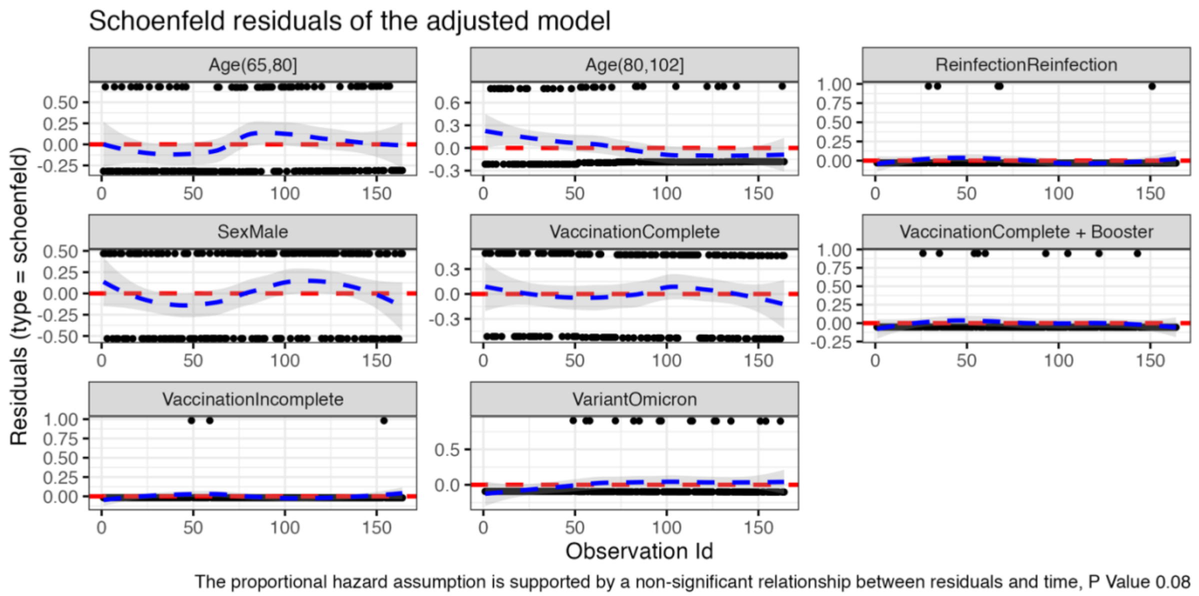


*Figure S4. Schoenfeld residuals of the adjusted model for hospitalization*

*Table S1. Odds Ratio for death in the adjusted model*

| | Odds ratio | Lower CI | Upper CI | P-value |
|---|---|---|---|---|
| **Omicron** | 0.140 | 0.001 | 1.122 | 0.069 |
| **Age** | | | | |
| (16,65] | | | | |
| (65,80] | 258.531 | 32.696 | 33367.876 | <0.001 |
| (80,102] | 4635.480 | 595.183 | 597754.200 | <0.001 |
| **Sex** | | | | |
| Male | 4.663 | 1.923 | 12.600 | <0.001 |
| **Reinfection** | | | | |
| Yes | 0.273 | 0.002 | 4.030 | 0.385 |
| **Vaccination** | | | | |
| Incomplete | 0.959 | 0.007 | 11.285 | 0.979 |
| Complete | 0.335 | 0.131 | 0.925 | 0.036 |
| Complete + Booster | 0.046 | 0.008 | 0.195 | <0.001 |
